# Evaluating the effect of mental health fine-tuning relative to other model characteristics on LLM safety performance

**DOI:** 10.64898/2026.01.02.25343289

**Authors:** Mark Kalinich, James Luccarelli, John Santa Maria, Gwydion Williams, Frank Moss, John Torous

## Abstract

Large language models (LLMs) are increasingly used in mental health applications, yet it remains unclear whether mental health–specific fine-tuning meaningfully improves safety-relevant performance beyond gains from model scale or architecture. We evaluated 127 publicly available open-source LLMs across three model families, multiple architecture generations, parameter scales (270M–70B), and fine-tuning strategies on three psychiatrist-reviewed synthetic classification tasks: suicidal ideation detection, identification of user requests for therapy, and detection of explicit therapy-like interactions in multi-turn conversations. Performance was summarized using F1 score, with multivariable regression and paired comparisons used to estimate independent effects of model characteristics. Across tasks, newer architectures and larger models consistently showed superior performance. General instruction tuning improved detection of therapy requests and engagement, whereas mental health–specific, medical, or safety fine-tuning conferred no consistent benefit and were sometimes associated with reduced performance. These findings suggest that baseline model capability is more consequential than domain-specific fine-tuning for certain safety-relevant mental health classification tasks, underscoring the importance of careful model selection and task-specific evaluation.

## Introduction

Large language models (LLMs) are increasingly being incorporated into mental health care. Recent systematic reviews of LLMs in mental health highlight rapid growth in applications spanning screening, conversational support, and other clinical workflows, while also emphasizing persistent risks and the need for rigorous evaluation approaches^1,2^. A primary strategy for adapting LLMs to specialized tasks is fine-tuning, in which an LLM that has been pre-trained on a large, diverse dataset (often non-clinical) is further trained on a focused domain-specific information (ie, clinical information)^3,4^. The goal of fine-tuning is to augment the model’s performance within a narrow performance space while retaining the knowledge and capabilities learned during pretraining.

Fine tuning has already shown impressive results. Models fine-tuned for instruction-following consistently outperform their non-fine-tuned counterparts on benchmarks assessing instruction adherence, reasoning, and human preference, leading to their widespread adoption for general-purpose, user-facing applications^5,6^. For instance, when blinded human evaluators were instructed to select their preferred model output to a given user input, the evaluators not only consistently preferred the outputs generated from 1.3-billion-parameter InstructGPT versus its non-fine-tuned counterpart, but still preferred InstructGPT’s outputs relative to the 175-billion-parameter base GPT-3 counterpart despite a >100-fold parameter difference^5^. Models that are fine-tuned using domain-specific data such as Meditron (on curated medical corpora) report ∼15% accuracy improvements over their base models across multiple medical benchmarks, and safety-tuned models like LLaMAGuard substantially outperform their base models on content risk detection^7,8^. Together, these examples demonstrate that fine-tuning, whether for general instruction following, specific domain knowledge, or explicit safety tasks often meaningfully improves model behavior relative to the base model.

But fine-tuning gains are not uniform and depend on baseline model scale, pre-training state, and evaluation scope relative to the specific fine-tuning task. Fine-tuning LLaMA-3 models yield substantial improvements for smaller models, but provides only limited marginal benefit for the 405B-parameter model on the massive multitask language understanding benchmark, which assesses knowledge and reasoning ability across 57 distinct subjects^9,10^. Similarly, fine-tuning models on medical corpora does not consistently improve clinical performance: for the same underlying models evaluated before and after biomedical fine-tuning, large models show little change across multiple clinical evaluation tasks, while smaller models actually underperform their pre-fine-tuned counterparts^11^. Fine-tuning that targets a specific behavior can also degrade performance on adjacent capabilities. In multimodal systems, continued fine-tuning of LLaVA-style models improves performance on the tuned dataset while reducing generalization and increasing hallucinations on closely related image classification datasets^12^. Fine-tuning can also introduce unexpected trade-offs, including increased over-refusal of benign requests following safety alignment^13,14^, and heightened sensitivity to prompt formulation or evaluation framing, limiting the stability and generalizability of observed gains^15,16^. Finally, pre-training and post-training can interact in non-intuitive ways. For example, recent peer-reviewed evidence on *catastrophic overtraining* shows that extending pre-training can make models harder to adapt, such that models fine-tuned from more extensively pre-trained checkpoints may perform worse than those derived from earlier checkpoints under identical fine-tuning procedures^17^.

Against this backdrop, the specific contribution of mental health–oriented fine-tuning to *safety classification* performance remains poorly characterized. Existing attempts to evaluate the performance of LLMs on mental health-related tasks, such as assessing the appropriateness of responses to individuals endorsing suicidal ideation, have demonstrated substantial variability across models^18,19^. However, few studies have explicitly quantified the relative contributions of domain-specific fine-tuning versus readily adjustable model characteristics (eg, parameter scale or model family) on concrete, safety-relevant classification tasks.

Establishing the marginal impact of mental health–specific fine-tuning in this setting can inform model selection and evaluation priorities for mental health–adjacent deployments, where fine-tuning is frequently assumed to provide meaningful benefit despite added cost, complexity, and potential unintended effects.

In this study, we evaluate a large set of publicly available open-source LLMs across multiple model families, parameter scales, and fine-tuning strategies on three safety-relevant classification tasks: suicidal ideation detection, identification of user requests for therapy, and recognition of therapy-like conversational interactions between a chatbot and user. Using standardized prompting and psychiatrist-reviewed benchmark data, we apply multivariable regression analyses to estimate the independent contributions of fine-tuning strategy, model scale, and model family to classification performance. Our goal is to clarify whether and under what conditions mental health–specific fine-tuning provides incremental benefit beyond gains attributable to newer, larger, or differently engineered base models.

## Results

**Figure 1.** highlights the significant heterogeneity in the number of models publicly available for a given fine-tuning type, model size, and architecture version, which limits the strength of comparative inferences, as the available models reflect a convenience sample rather than a controlled experimental design.

**Figure 1.**
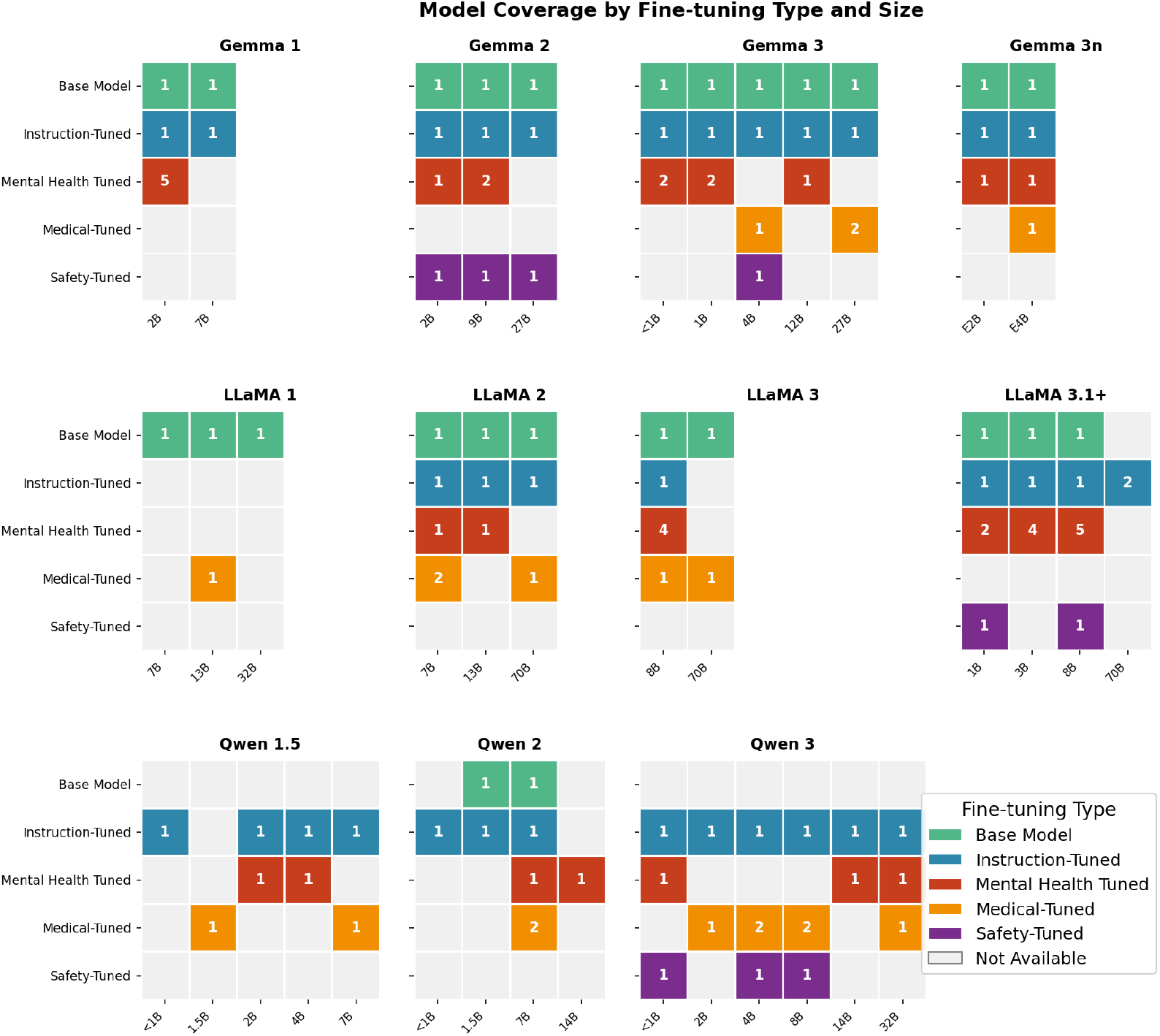
Available LLMs by model family, model architecture version (as defined by the model creator), number of parameters, and fine-tune method. Numbers represent the number of available models with a given type of fine-tune and parameter count for a given model family and architecture.

In order to discern general trends in the dataset, F1 score for each of the three prediction tasks was plotted against model parameter size and segmented by model family (Figure 2). Linear regression on log-transformed parameter counts revealed significant positive associations for five of nine family-task combinations after Bonferroni correction for multiple hypothesis testing. Gemma models showed the most consistent scaling benefits, with significant improvements across all three classification tasks: suicidal ideation (R^2^ = 0.28, p_adjusted_ < 0.01), therapy request (R^2^ = 0.23, p_adjusted_ < 0.01), and therapy engagement (R^2^ = 0.27, p_adjusted_ < 0.01). Qwen models exhibited the strongest parameter scaling effect for suicidal ideation detection (R^2^ = 0.32, p_adjusted_ < 0.01), while LLaMA models showed significant improvement only for therapy request classification (R^2^ = 0.21, p_adjusted_ < 0.05). The remaining four family-task combinations (LLaMA suicidal ideation and therapy engagement, Qwen therapy request and therapy engagement) showed positive but non-significant trends. Detailed performance characteristics for each classification task can be found in **Figures S1-9**.

**Figure 2.**
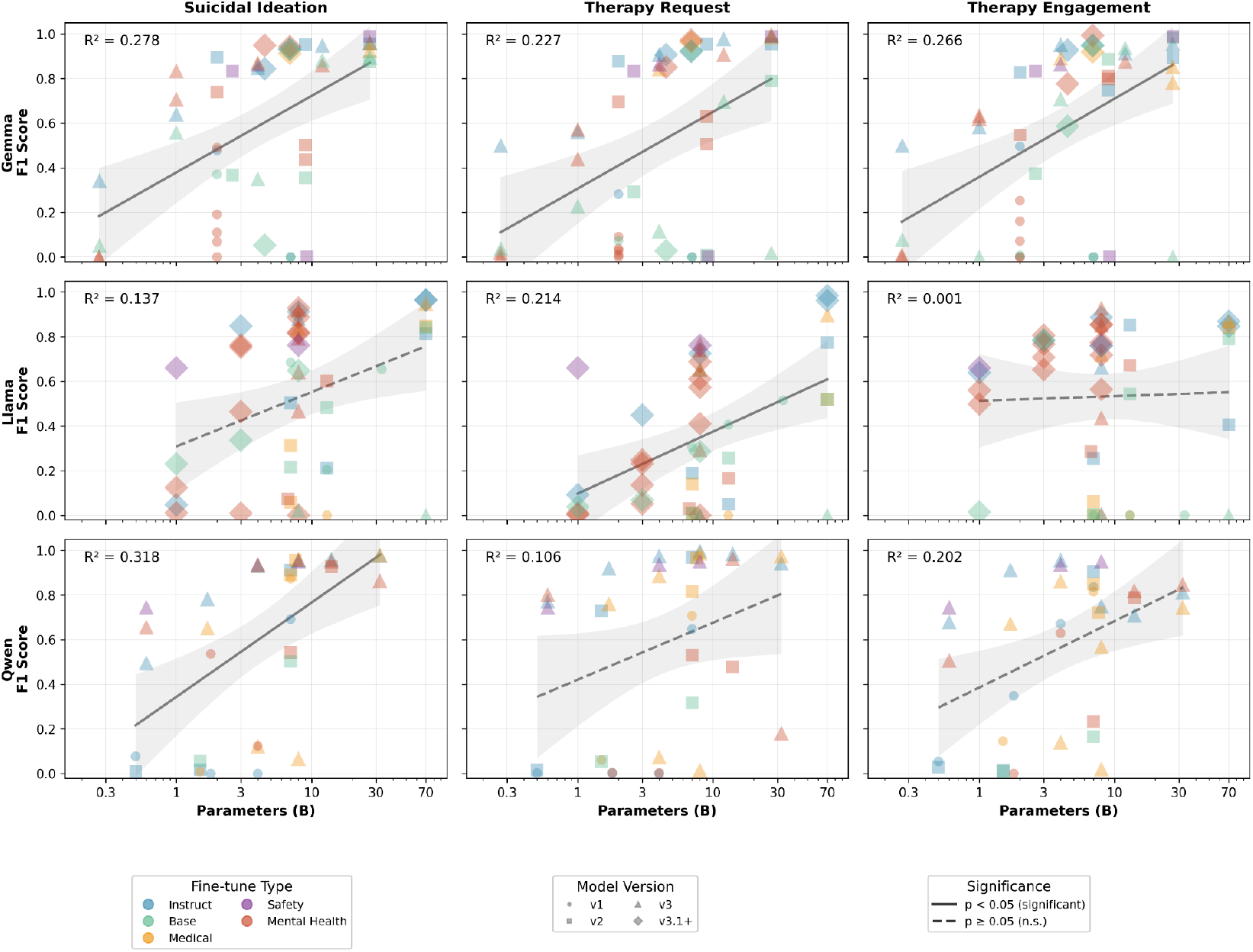
Model performance (F1 score) versus parameter count across classification task, fine tuning approach, and model version

To quantitatively estimate the relative independent contributions of model family, model version, fine-tuning, and model parameter size to model performance, multivariable linear regression was performed (**Table 1**). Newer model architecture versions demonstrated a statistically significant association with model performance (F1 score) across all tasks. Although Version 2 models only showed a significant difference relative to Version 1 for therapy request (β = 0.27, p_adjusted_ <.05) and therapy engagement detection (β = 0.28, p_adjusted_ <.05), Version 3 models showed significantly higher F1 scores for suicidal ideation detection (β = 0.36, p_adjusted_ <.001), therapy request detection (β = 0.38, p_adjusted_ <.001), and therapy engagement detection (β = 0.40, p_adjusted_ <.001) relative to Version 1. Version 4 models demonstrated even larger improvements (suicidal ideation: β = 0.52, p <.01; therapy request: β = 0.62, p <.001; therapy engagement: β = 0.65, p_adjusted_ <.001). The number of parameters in the model was significant for SI and therapy-request detection, but not for therapy-engagement detection.

**Table 1.** Multivariable regression of F1 score against model architecture version, parameter size, fine-tune type, and model family.

| Variable | Suicidal Ideation |  | Therapy Request |  | Therapy Engagement |  |
| --- | --- | --- | --- | --- | --- | --- |
| | $\beta$ | 95% CI | $\beta$ | 95% CI | $\beta$ | 95% CI |
| Intercept | 0.118 | [-0.155, 0.392] | -0.049 | [-0.325, 0.227] | 0.016 | [-0.221, 0.253] |
| Version: 2 | 0.229 | [-0.046, 0.505] | 0.266* | [0.003, 0.529] | 0.285* | [0.032, 0.539] |
| Version: 3 | 0.360*** | [0.113, 0.607] | 0.378*** | [0.136, 0.620] | 0.396*** | [0.183, 0.608] |
| Version: 4 | 0.515** | [0.112, 0.918] | 0.620*** | [0.226, 1.015] | 0.654*** | [0.413, 0.896] |
| Parameter Size (B) | 0.008* | [0.001, 0.014] | 0.007** | [0.002, 0.012] | 0.004 | [-0.002, 0.010] |
| Fine-Tune Type: Instruction-Tuned | 0.184 | [-0.079, 0.447] | 0.347*** | [0.104, 0.591] | 0.303** | [0.046, 0.560] |
| Fine-Tune Type: Mental Health Tuned | 0.139 | [-0.103, 0.381] | 0.145 | [-0.084, 0.374] | 0.219 | [-0.024, 0.462] |
| Fine-Tune Type: Medical-Tuned | 0.169 | [-0.139, 0.477] | 0.295* | [0.005, 0.585] | 0.190 | [-0.116, 0.497] |
| Fine-Tune Type: Safety-Tuned | 0.286 | [-0.067, 0.639] | 0.446** | [0.091, 0.801] | 0.360 | [-0.021, 0.741] |
| Family: LLaMA | -0.136 | [-0.333, 0.061] | -0.218* | [-0.410, -0.025] | -0.073 | [-0.270, 0.124] |
| Family: Qwen | 0.015 | [-0.185, 0.214] | 0.031 | [-0.166, 0.228] | 0.002 | [-0.201, 0.205] |
| **R<sup>2</sup>** | **0.335** |  | **0.486** |  | **0.374** |  |
| **Adj R<sup>2</sup>** | **0.278** |  | **0.441** |  | **0.320** |  |
| **N** | **127** |  | **127** |  | **127** |  |
| **Significance levels (Bonferroni-corrected):** *p<0.05; **p<0.01; ***p<0.001<br>**Multiple testing correction:** Bonferroni correction applied to p-values AND confidence intervals<br>**Confidence intervals:** Bonferroni-adjusted ($\alpha = 0.05/n_{\text{tests}}$)<br>**Reference categories:** Family=Gemma, Version=1, Fine-Tune Type=Base Model<br>**Standard errors:** HC3 heteroscedasticity-consistent (robust) |  |  |  |  |  |  |

Relative to the base, pre-trained models, mental health fine-tuning did not demonstrate statistically significant improvements on any of the three tasks. Models that were post-trained for general instruction following were associated with significantly higher F1 scores for therapy request (β = 0.35, p_adjusted_ <.001) and therapy engagement (β = 0.30, p_adjusted_ <.01) tasks, but not for suicidal ideation detection. Safety-tuned models showed significant improvements only for therapy request detection (β = 0.45, p_adjusted_ <.01). Model family (LLaMA vs. Gemma, Qwen vs. Gemma) generally did not significantly predict performance, with the exception of LLaMA showing lower therapy request F1 scores compared to Gemma (β = -0.22, p <.05). Overall, these models explained 34–49% of variance in F1 scores (R^2^ = 0.34–0.49), with therapy request prediction showing the strongest model fit.

To directly isolate the effect of mental health–specific fine-tuning from differences attributable to model family, architecture, or scale, we next restricted the analysis to fine-tuned models with clearly identified and publicly available pre-fine-tuned model counterparts, which are listed in **Table S2**. This paired design allowed within-model comparisons under identical prompting and evaluation conditions, minimizing confounding from architectural generation and parameter count that characterize the broader model set. Note that fine-tuned models could originate from either base models or be domain-specific fine-tuning on top of a model already fine-tuned for instruction-following.

Across all three safety-relevant classification tasks and all evaluated model families, mental health–specific fine-tuning did not yield statistically significant improvements in performance relative to the exact pretrained base models from which those fine-tuned variants were derived (**Figure 3A**). In contrast, several mental health–fine-tuned models demonstrated statistically significant *decreases* in mean Δ F1 compared with their corresponding base models, including Gemma (Δ F1 = -0.19, p_adjusted_ < 0.01) and LLaMA (Δ F1 = -0.17; p_adjusted_ < 0.05) models on therapy-request detection, and Gemma (median Δ F1 = -0.17, p_adjusted_ < 0.05) models on therapy-engagement classification (two-sided paired *t* tests, Bonferroni-corrected). These effects were observed despite identical prompting, evaluation conditions, and shared model backbones for each base–fine-tuned pair.

**Figure 3.**
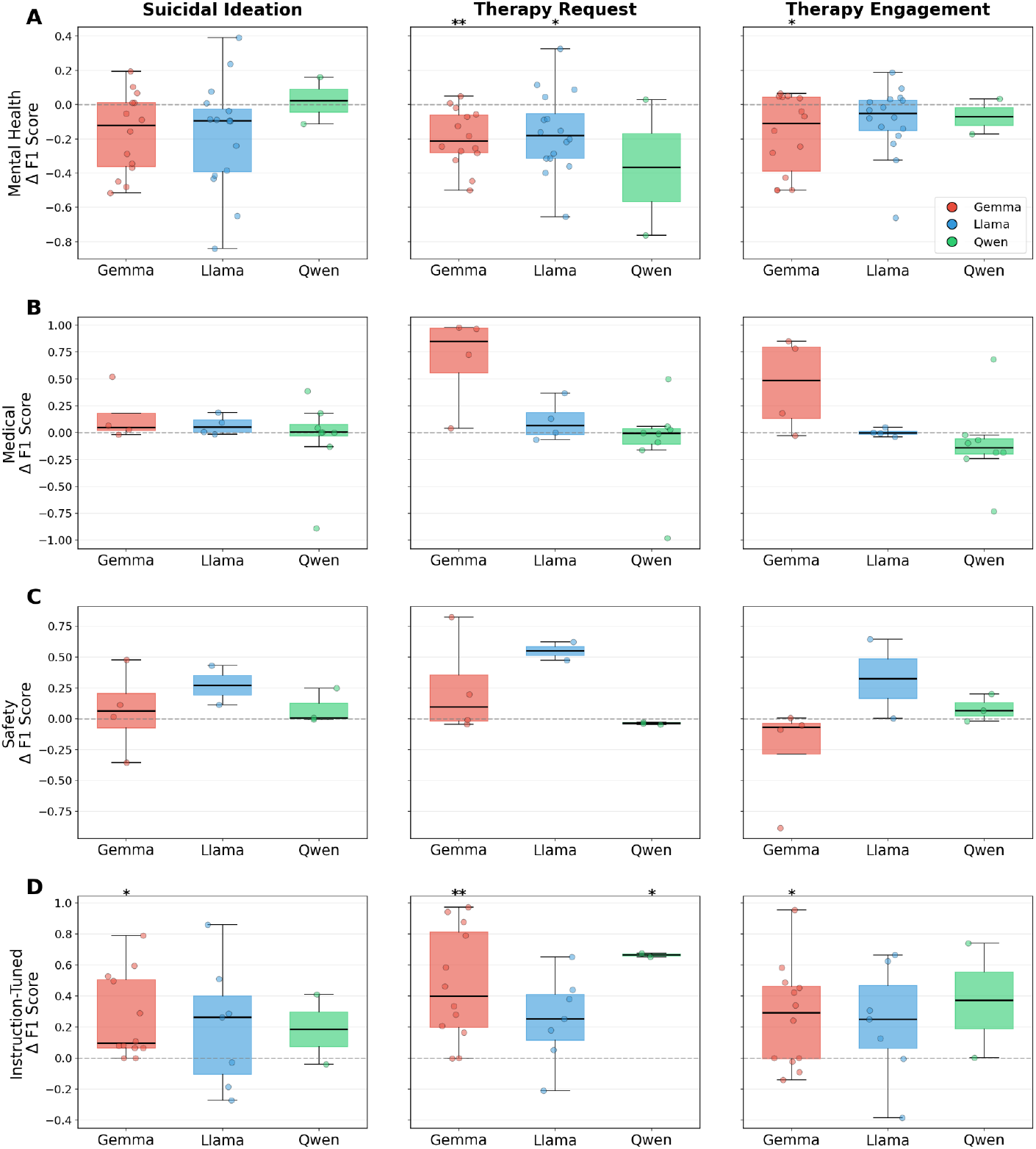
Change in performance between fine-tuned models and their base counterparts. A) Mental health; B) general medical activities, C) unsafe content detection D) general instruction-tuned models. Legend: *p_adjusted_ < 0.05, **p_adjusted_ < 0.01, ***p_adjusted_ < 0.001.

Models fine-tuned on medical corpora yielded no statistically significant differences in performance compared with base models across any task or model family, although Gemma models showed large mean Δ F1’s for therapy request (0.68) and therapy engagement (0.45) detection (**Figure 3B**). Similar null results were observed for safety-focused fine-tunes (**Figure 3C**): despite the explicit harm-detection objectives of models such as ShieldGemma, LLaMA Guard, and Qwen Guard, no family demonstrated significant improvements on any of the three safety-relevant classification tasks relative to their pre-fine-tuned models. These findings suggest that, in their default configurations, neither medical nor safety fine-tuning conferred reliable benefits on this set of mental health safety classification tasks.

In contrast to domain-specific fine-tuning, general fine-tuning for instruction-following demonstrated statistically significant improvements in classification performance for at least one model family in all three classification tasks (**Figure 3D**). Gemma instruction-tuned models significantly outperformed their pretrained counterparts across all three tasks: suicidal ideation detection (median Δ F1 = 0.26, p_adjusted_ < 0.05), therapy-request classification (median Δ F1 = 0.47 p_adjusted_ < 0.01), and therapy-engagement detection (Δ F1 = 0.27, p_adjusted_ < 0.05). Qwen instruction-tuned models also showed significant improvement on therapy-request detection (Δ F1 = 0.66, p_adjusted_ < 0.05). Notably, the overall direction of effect was overwhelmingly positive: 47 of 63 instruction-tuned model pairs showed improved performance relative to their pretrained bases.

## Discussion

In this large-scale evaluation of open-source large language models across three safety-relevant mental health classification tasks, the most consistent determinants of performance were model generation (architecture/version) and parameter scale. Across model families, newer architecture generations were associated with reliably higher classification performance, and larger models generally performed better—particularly for suicidal ideation and therapy request detection—though effects were not uniform across tasks. Against this backdrop, we also observed that general instruction-tuned variants tended to perform more consistently than pretrained base models, reflecting improved instruction adherence and structured output compliance under standardized prompting.

In contrast, mental health–specific fine-tuning did not yield reliable incremental benefit once model version, size, and general instruction-tuning were accounted for. In paired comparisons restricted to fine-tuned models with clearly identified base counterparts, mental health–fine-tuned variants did not significantly outperform their base models under identical prompts and evaluation conditions, and several showed statistically significant performance decrements. Consistent with this, multivariable regression suggested that the magnitude and reliability of architecture- and scale-associated gains exceeded those attributable to fine-tuning category, indicating that baseline model capability remains the primary driver of safety classification behavior in this setting. Because deployment-grade safety classifiers require both *correct discrimination* and *reliable structured outputs*, we interpret performance as reflecting a composite of semantic detection and instruction/format compliance.

These findings refine common assumptions in mental health–adjacent LLM development. Fine-tuning is often treated as the main lever for improving clinically relevant behavior, but our results suggest that selecting a newer-generation model and appropriate scale may matter more than applying mental health–specific fine-tuning to an older or weaker backbone. These results indicate that general-purpose instruction tuning, which emphasizes instruction-following and conversational coherence, may be more beneficial for zero-shot safety classification than fine-tuning on domain-specific corpora.

It is also important to emphasize that model-level behavior does not fully determine the safety properties of deployed LLM-based products. In real-world applications, language models are embedded within broader systems that include prompt orchestration, routing and gating logic, retrieval-augmented generation, and dedicated safeguard or moderation components. These system-level controls can substantially shape how safety-relevant content is handled, often independently of the underlying model’s intrinsic classification behavior. As a result, performance differences observed at the model level may not translate directly to deployed settings, and meaningful evaluation of clinical risk will require more sophisticated, multidimensional, and longitudinal assessment of the integrated system rather than the language model in isolation.

This study has several limitations. First, these findings are based on simulated clinical interactions. Validation using real-world data collected across diverse clinical scenarios will be necessary to assess generalizability, confirm applicability to any intended user population, and identify context-dependent performance differences. Second, the safety-related tasks evaluated here represent a deliberately narrow operationalization intended to support relative comparisons of fine-tuning effects rather than a comprehensive assessment of clinical safety. The use of structured prompts and constrained outputs facilitates standardized benchmarking but does not reflect typical clinical interactions, and performance on these tasks should not be interpreted as evidence of real-world safety. Third, the evaluated models comprise a convenience sample of readily accessible open-source GGUF checkpoints, providing insight into the current state of open-source tooling but resulting in heterogeneous training quality and uneven coverage across model families, versions, and parameter scales. Fourth, all evaluations were conducted in English and were not assessed across other languages or cultural contexts, limiting generalizability. Fifth, multiple fine-tuned models share the same base model, which may violate the statistical assumption of independence and inflate our significance calculations. Finally, the multivariable regression analyses are descriptive and not intended to support causal inference. Aggregating across heterogeneous model families and fine-tuning approaches necessarily conflates multiple design choices, and the results should be interpreted as summarizing observed associations under standardized evaluation conditions.

In summary, across 127 open-source LLMs evaluated on three safety-relevant mental health classification tasks, performance differences were most strongly associated with model generation, model scale, and general instruction tuning in classifier-ready behavior under structured prompting. In contrast, the publicly available mental health–, medical-, and safety-tuned variants evaluated here did not demonstrate reliable incremental improvements over their corresponding base models, and several mental health fine-tunes showed performance decrements, underscoring that “mental health tuning” optimized for therapeutic dialogue does not necessarily translate into better safety classification. These findings suggest that, for developers deploying LLMs in mental health–adjacent settings, careful selection of a strong, modern backbone model may be more consequential for safety-relevant detection than applying domain-oriented fine-tuning alone. As more capable foundation models continue to emerge, future work should test whether targeted, task-matched fine-tuning on clinically grounded data can yield robust gains beyond those attributable to architecture and scale, and should validate these findings using real-world clinical data and standardized evaluation frameworks before any clinical deployment

## Methods

### Synthetic dataset construction and clinical validation

Synthetic data were generated using Gemini 2.5 Pro to support three safety-relevant classification tasks, following our previously described approach^20^. Briefly, Gemini was instructed to produce diverse, clinically plausible user and chatbot utterances with variation in tone, affect, formality, and sentence structure. This included 1,000 statements for suicidal ideation detection, 1,200 statements for therapy-request detection, and 450 four-turn dialogues spanning clearly therapeutic, clearly non-therapeutic, and ambiguous interactions. Two psychiatrists independently reviewed all outputs: the first clinician approved, modified, or removed statements; the second clinician validated the approved or modified set. Each category was down-sampled to ensure class balance prior to model testing.

### Model selection

We evaluated 127 publicly available open-source large language models spanning three major model families: Gemma-derived, LLaMA-derived, and Qwen-derived models. Multiple architecture generations were represented within each family, including Gemma (v1, v2, v3, MedGemma, and Gemma 3N), LLaMA (v1, v2, v3, 3.1, 3.2, and 3.3), and Qwen (Qwen1.5, Qwen2, and Qwen3). Model sizes ranged from 270M to 70B parameters. Models reflected several fine-tuning categories, including base (pretrained) models, instruction-tuned general-purpose models, general medical models trained on biomedical or clinical corpora (including multimodal clinical models), mental health–specific models fine-tuned on counseling or mood-related data, and safety-aligned models designed for content filtering or risk classification (**Figure 1**). All models were downloaded directly through LM Studio with GGUF-formatted weights, using between 4- and 8-bit quantization. Evaluated models and configurations are listed in **Table S1**.

### Model evaluation

Model evaluation followed a previously described framework^20^. All models were executed locally in LM Studio using fixed decoding parameters (temperature = 0.0, top_p = 1.0, max_tokens = 256) to ensure deterministic outputs. Structured prompts corresponding to each task required models to produce standardized JSON-formatted labels. Model responses were obtained via LM Studio’s local inference API and cached in a SQLite database using composite hash keys derived from the model, prompt, and input data to enable reproducible evaluation. Binary classification performance metrics (sensitivity, specificity, accuracy, and F1 score) were computed by parsing model outputs and comparing predicted labels against clinically adjudicated ground truth labels. Safety-specialized models (ShieldGemma, LLaMA Guard, and Qwen Guard) were evaluated using their native output formats rather than the standardized JSON schema. ShieldGemma models were distributed with a Jinja2 template incompatible with LM Studio; this template was replaced with a standard Gemma chat template to enable evaluation.

### Statistical Analysis

To assess the impact of model characteristics on classification performance, we conducted multivariable linear regression analyses for each of three mental health classification tasks (suicidal ideation detection, therapy request detection, and therapy engagement detection), with F1 score as the dependent variable. Independent variables included model family (categorical: Gemma, LLaMA, Qwen), architecture version (ordinal: 1, 2, 3, 4), parameter size (continuous, in billions), and fine-tune type (categorical: Base Model, Instruction-Tuned, Medical-Tuned, Mental Health Tuned, Safety-Tuned), with all 127 enabled models included in the analysis; HC3 heteroscedasticity-consistent (robust) standard errors were used to account for potential heterogeneity in error variance across model types.

To directly estimate the effect of mental health–specific fine-tuning, independent of model architecture and scale, we conducted a paired-comparison analysis restricted to fine-tuned models with clearly identified, and publicly available base models (**Table S2**). For each eligible mental health–fine-tuned model, we evaluated the corresponding base model using identical prompts and benchmark data, and computed within-pair differences in performance metrics. These paired differences were then analyzed using paired *t* tests to assess whether mental health–specific fine-tuning was associated with statistically significant changes in classification performance relative to the base model.

### Computational environment

All models were evaluated locally on a dedicated workstation using pre-downloaded weights. Inference was performed on a Dell Pro Max Tower T2 workstation equipped with an NVIDIA RTX PRO 6000 Blackwell GPU (96 GB VRAM), an Intel Core Ultra 9 285K CPU (24 cores, 24 threads), and 128 GB of system memory. The system ran Red Hat Enterprise Linux 9.7 (kernel 5.14.0) with Python 3.9.23 in a virtual environment. Data processing, metric computation, and visualization were performed using pandas 2.2.3, NumPy 1.26.4, scikit-learn 1.5.2, matplotlib 3.9.2, and seaborn 0.13.2. Models were executed using LM Studio version 0.3.31 (Build 7) with GGUF-formatted checkpoints (see **Table S1** for model details). All data generated in the present study are included in the Supplementary Materials. Data processing and analysis scripts are available at: https://github.com/markkalinich/mental-health-finetune-analysis

## Supporting information

All model inputs and outputs for all experiments

## Data availability statement

All data produced in the present work are contained in the manuscript and supplementary materials.

## Code availability statements

Data processing scripts are available at https://github.com/markkalinich/mental-health-finetune-analysis.

## Acknowledgment

MK and JT receive funding from Harvard Medical School and the BIDMC Department of Psychiatry. JL receives funding from NIMH, Harvard Medical School, the Rappaport Foundation, the Foundation for Prader-Willi Research, and the American Academy of Child and Adolescent Psychiatry. JSM is employed by Parabilis Medicines.

While preparing this work, the authors used Claude Sonnet and Opus v4.5 (via Cursor) to architect and perform the computational experiments designed above. GPT-5.1-2 was used during the editing of this manuscript, and to aggregate data about the models used. After using these tools, the authors reviewed and edited the content as needed and take full responsibility for the content of the published article.

## Author contributions

MK and JT conceived the study. MK conducted the experiments and analyses. JT and MK co-wrote the first draft of the manuscript. JSM, FM, JL, and GW contributed to the study design and edited and revised the manuscript. All authors approved the final manuscript.

