## Supplementary Figures for "Evaluating the effect of mental health fine-tuning relative to other model characteristics on LLM safety performance"

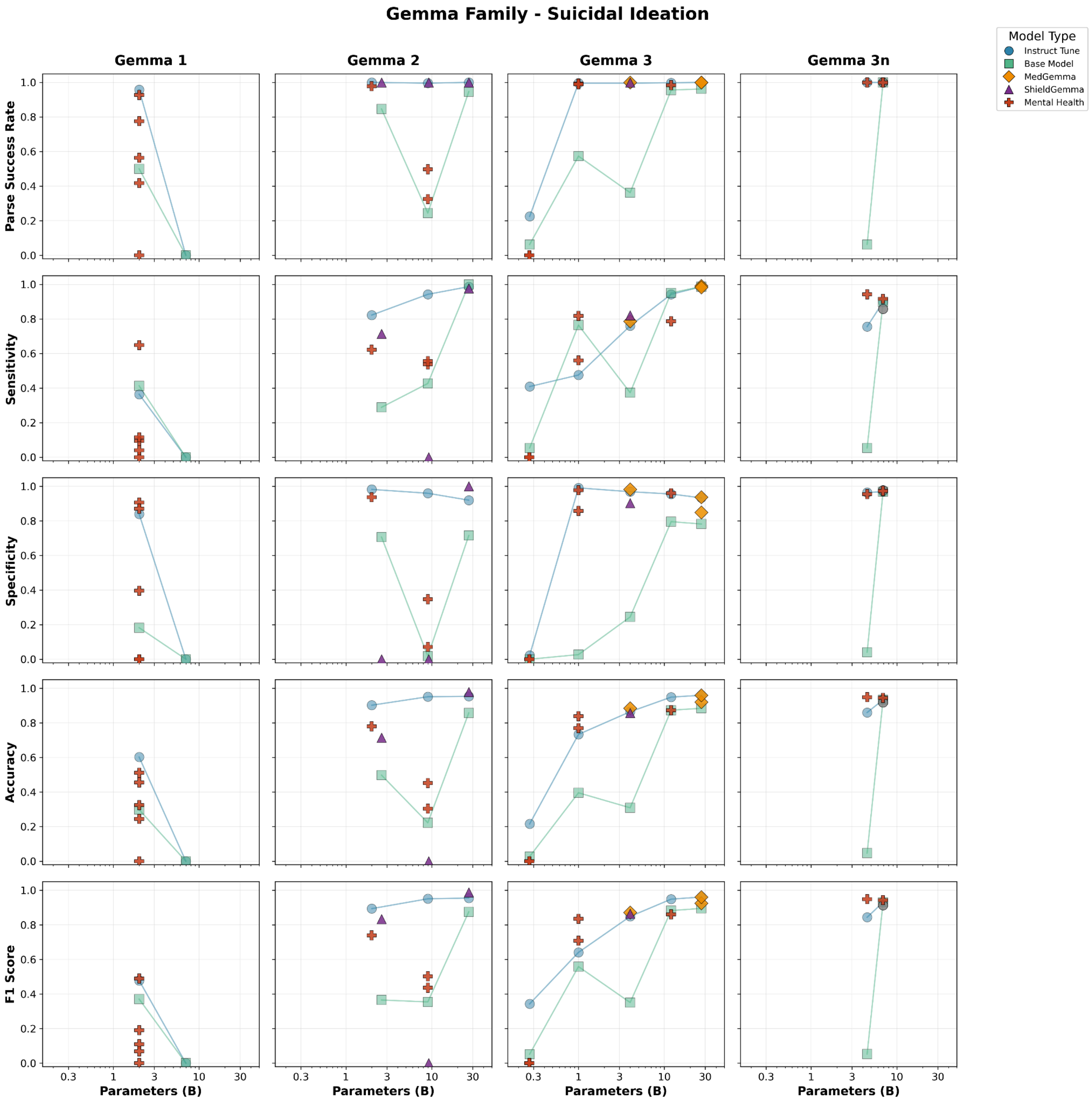


**Figure S1:** Detailed performance characteristics for suicidal ideation detection task for Gemma Models

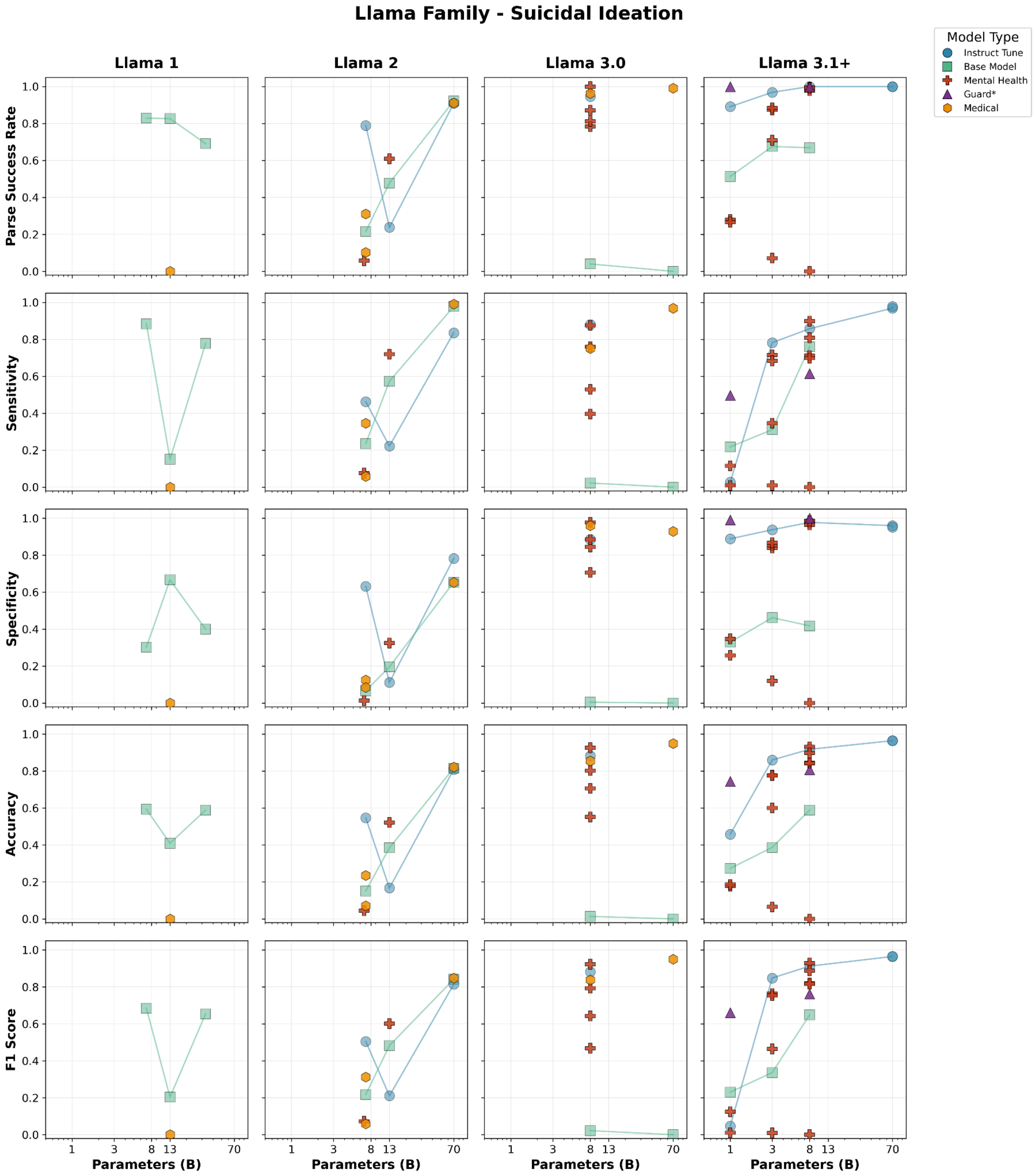


**Figure S2:** Detailed performance characteristics for suicidal ideation detection task for LLaMA Models

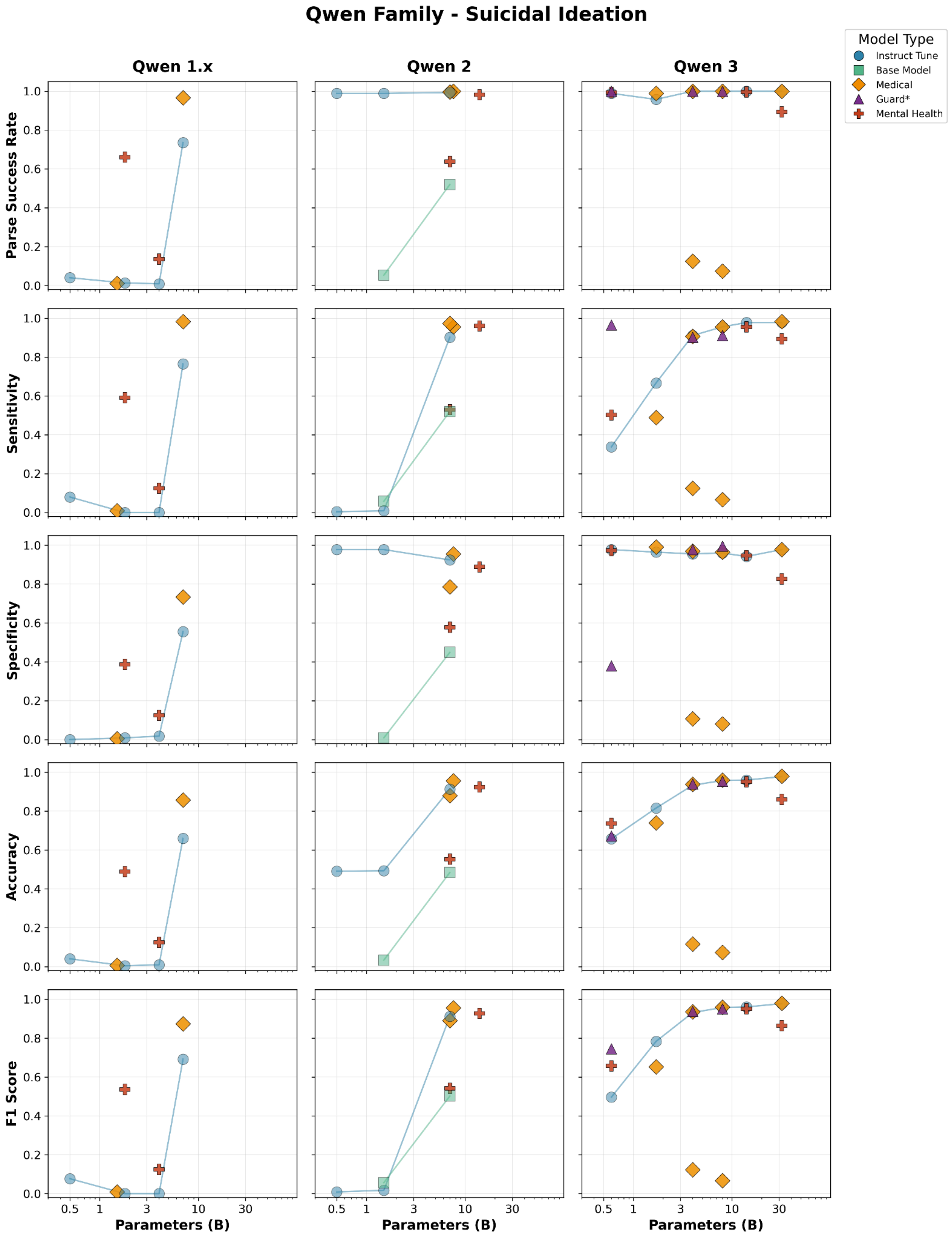


**Figure S3:** Detailed performance characteristics for suicidal ideation detection task for Qwen Models

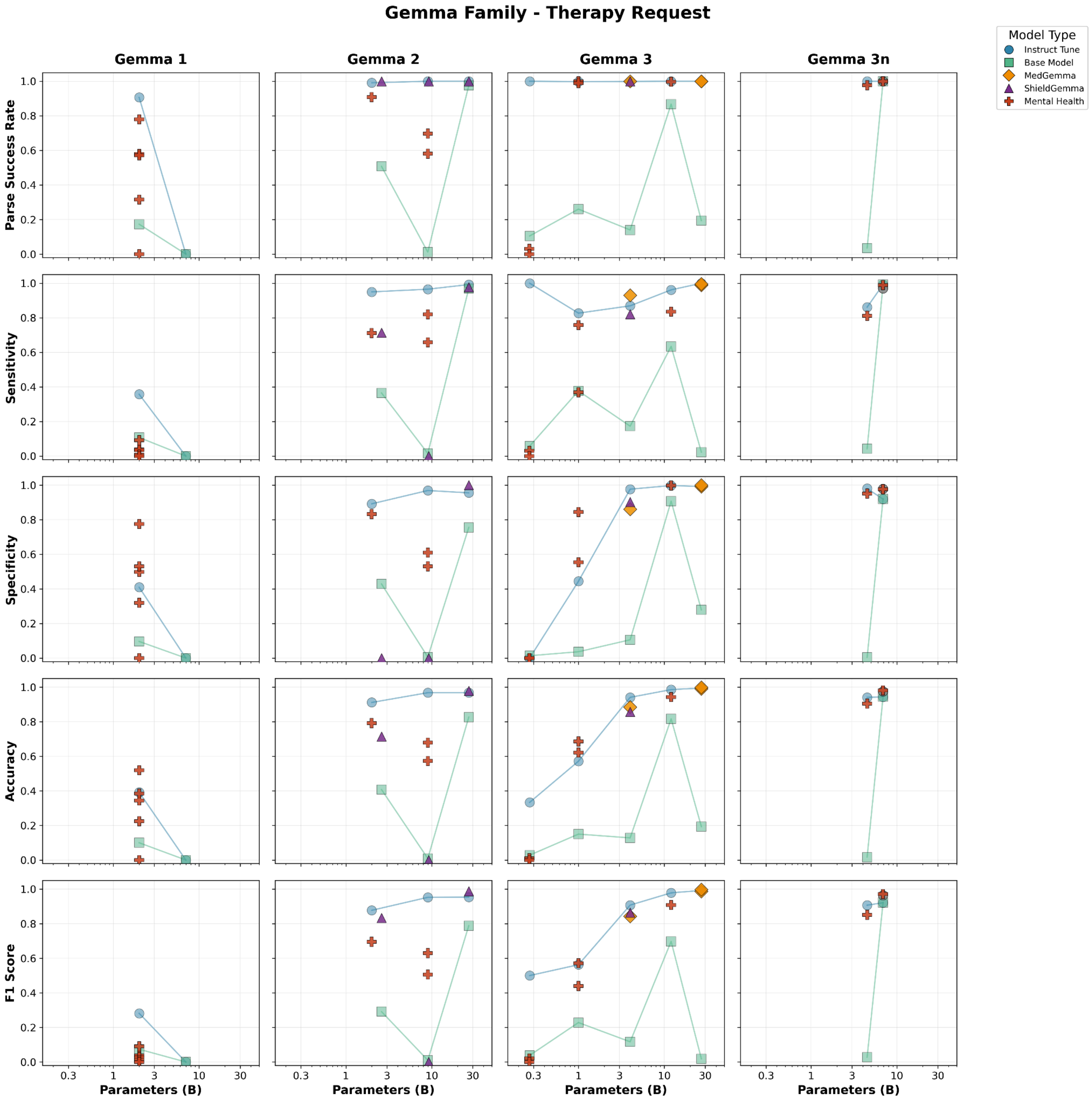


**Figure S4:** Detailed performance characteristics for therapy request detection task for Gemma Models

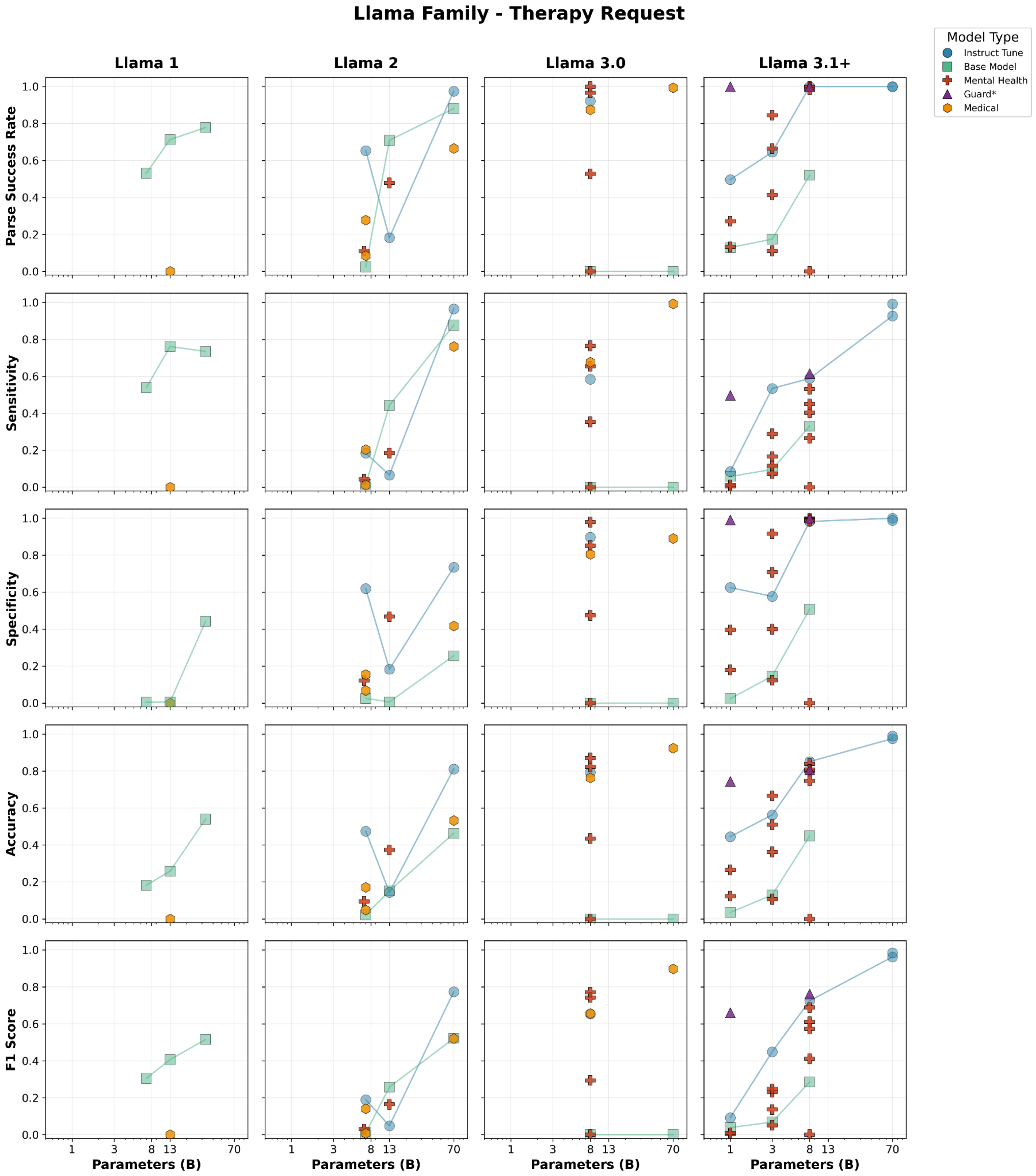


**Figure S5:** Detailed performance characteristics for therapy request detection task for LLaMA Models

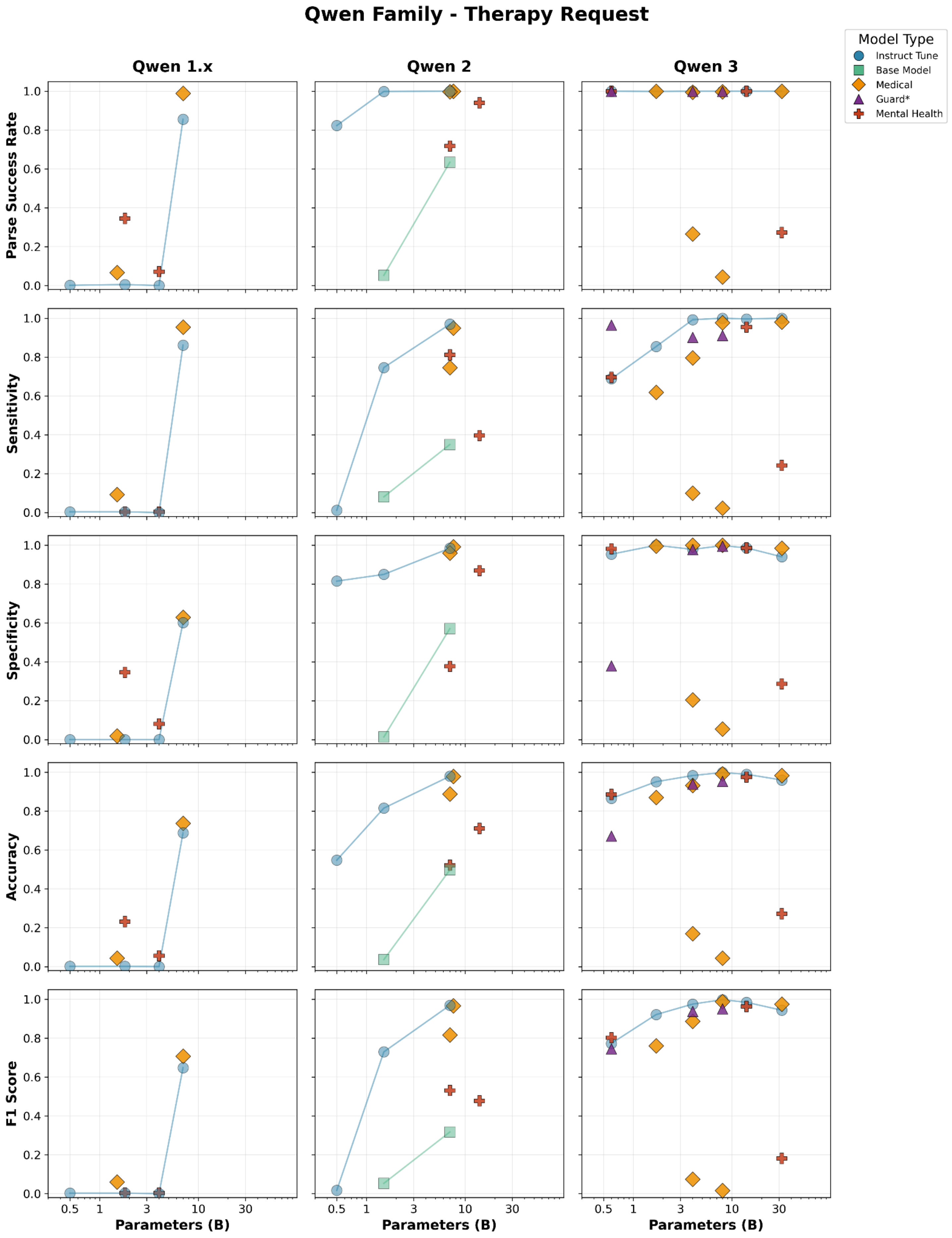


**Figure S6:** Detailed performance characteristics for therapy request detection task for Qwen Models

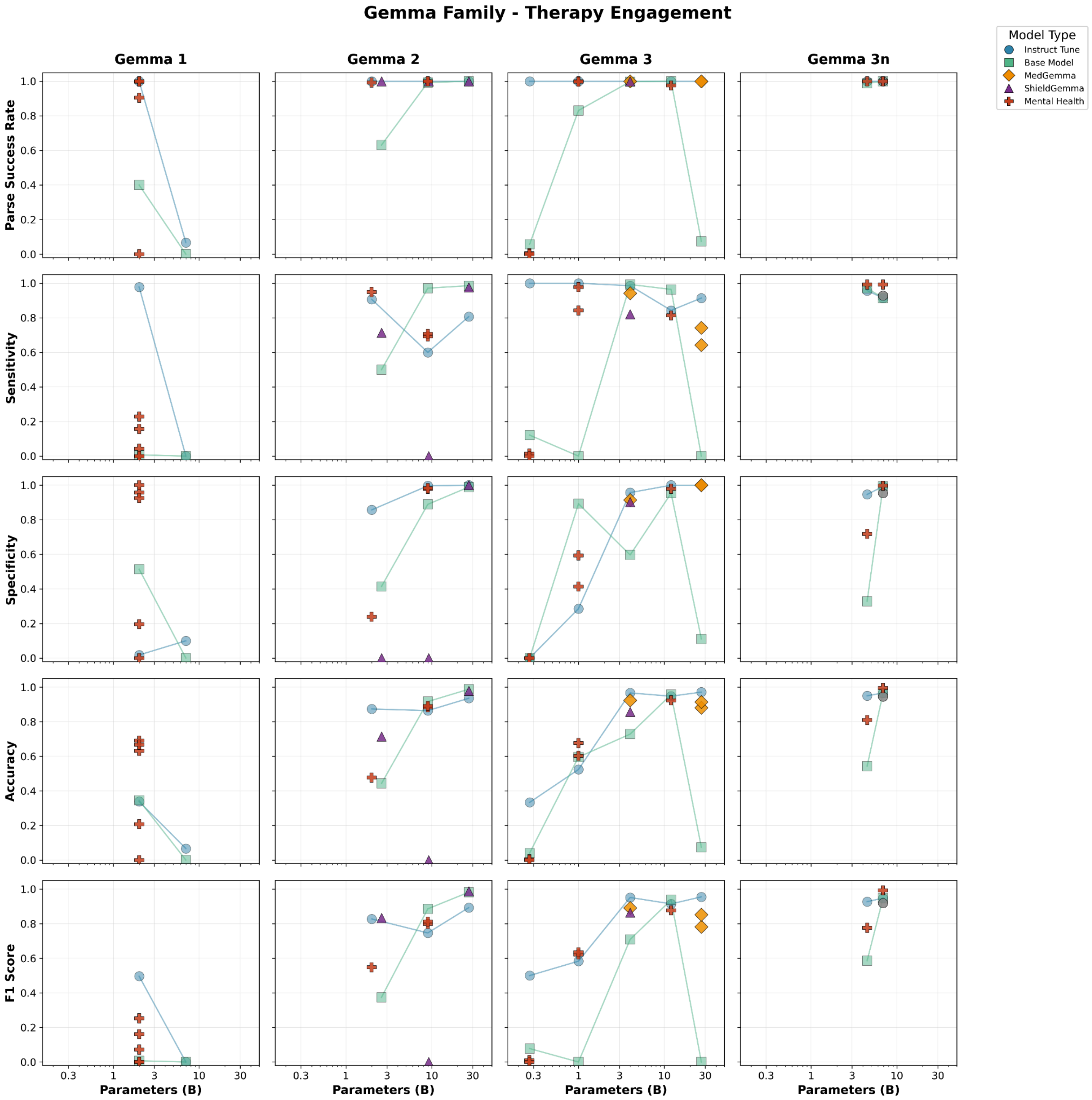


**Figure S7:** Detailed performance characteristics for therapy engagement detection task for Gemma Models

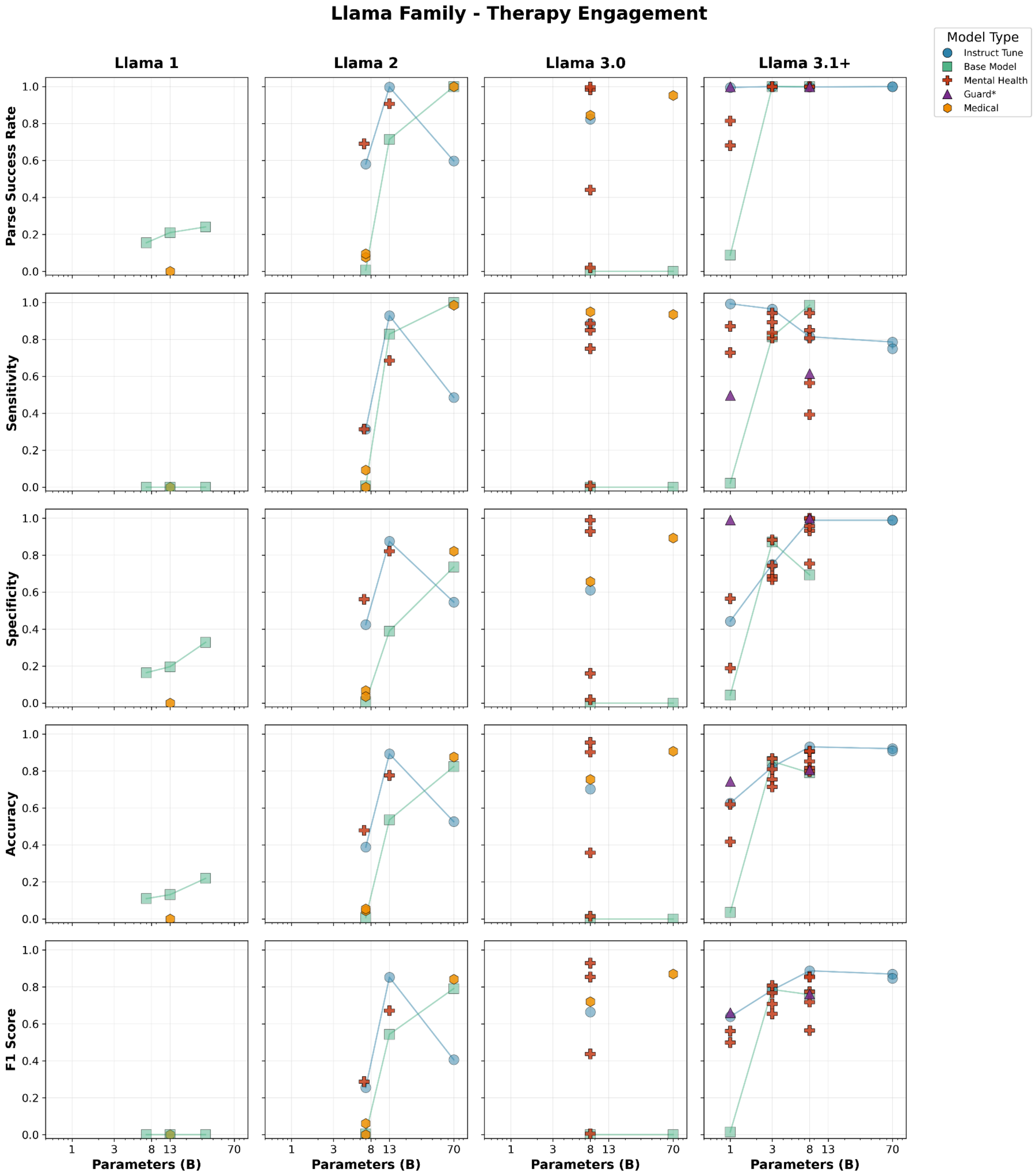


**Figure S8:** Detailed performance characteristics for therapy engagement detection task for LLaMA Models

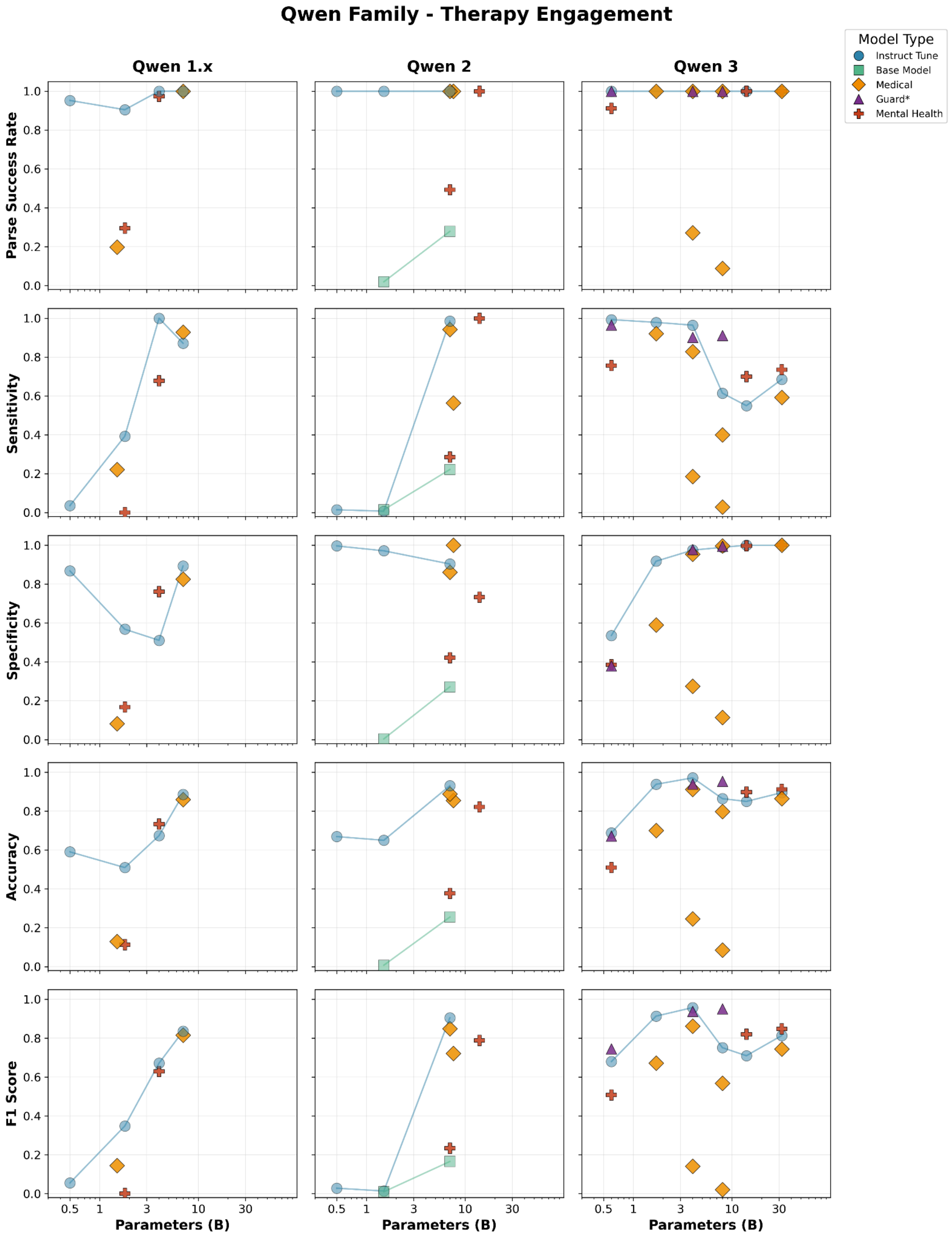


**Figure S9:** Detailed performance characteristics for therapy engagement detection task for Qwen Models
